# Analyzing wav2vec embedding in Parkinson’s disease speech: A study on cross-database classification and regression tasks

**DOI:** 10.1101/2024.04.10.24305599

**Authors:** Ondrej Klempir, Radim Krupicka

## Abstract

Advancements in deep learning speech representations have facilitated the effective use of extensive datasets comprised of unlabeled speech signals, and have achieved success in modeling tasks associated with Parkinson’s disease (PD) with minimal annotated data. This study focuses on PD non-fine-tuned wav2vec 1.0 architecture. Utilizing features derived from wav2vec embedding, we develop machine learning models tailored for clinically relevant PD speech diagnosis tasks, such as cross-database classification and regression to predict demographic and articulation characteristics, for instance, modeling the subjects’ age and number of characters per second. The primary aim is to conduct feature importance analysis on both classification and regression tasks, investigating whether latent discrete speech representations in PD are shared across models, particularly for related tasks. The proposed wav2vec-based models were evaluated on PD versus healthy controls using three multi-language-task PD datasets. Results indicated that wav2vec accurately detected PD based on speech, outperforming feature extraction using mel-frequency cepstral coefficients in the proposed cross-database scenarios. Furthermore, wav2vec proved effective in regression, modeling various quantitative speech characteristics related to intelligibility and aging. Subsequent analysis of important features, obtained using scikit-learn feature importance built-in tools and the Shapley additive explanations method, examined the presence of significant overlaps between classification and regression models. The feature importance experiments discovered shared features across trained models, with increased sharing for related tasks, further suggesting that wav2vec contributes to improved generalizability. In conclusion, the study proposes wav2vec embedding as a promising step toward a speech-based universal model to assist in the evaluation of PD.

## 1 Introduction

In recent years, the application of deep neural networks has revolutionized the field of medical research, offering innovative solutions to complex machine-learning challenges [1]. These networks have demonstrated state-of-the-art advancements across various medical domains [2, 3]. The integration of deep learning (DL) models and wearable technology has emerged as a promising solution for the diagnosis and subsequent monitoring in medical contexts, particularly for Parkinson’s disease (PD) [4–6]. DL has proliferated in speech processing and language understanding. In addition to traditional methods based on calculating features from audio recordings [7], highly performing speech representations like wav2vec [8, 9] have emerged, leading to improved performance in various tasks such as automatic speech recognition [10] or emotion analysis from voice [11]. In the case of speech disorders, this includes specific applications for dysarthric speech classification [12]. An advanced application involves attempts to decode speech from brain activity [13]. While wav2vec has proven its capability to generalize across languages [14], an open question persists concerning PD, i.e. the determination of whether a particular subset of wav2vec features exhibits generalization across various diagnostic speech tasks.

PD stands as the second most prevalent degenerative disorder of the central nervous system, following Alzheimer’s disease. It affects about 1 in 100 people 65 years of age or older [15]. Its etiology is closely linked to the degeneration of dopamine-forming nerve cells within the substantia nigra, a critical component of the basal ganglia complex [16]. Speech, an integral indicator of motor functions and movement coordination, has proven to be exceptionally sensitive to central nervous system involvement [17]. Major movement symptoms of PD, including tremor, stiffness, slowing, and disturbances in posture and gait, only manifest when a substantial portion of brain cells are affected. In contrast, alterations in speech can occur significantly earlier, up to a decade prior to a formal diagnosis [18]. Recognizing speech impairment in the early stages of PD is crucial for a timely prognosis and the implementation of appropriate therapeutic measures [19].

This paper focuses on analyzing the feature importance of wav2vec 1.0 in PD speech and investigates whether the latent discrete speech representations are shared across tasks, with increased sharing for related tasks. The study explores sets of features contributing across various datasets (participants rhythmically repeat syllable /pa/, Italian dataset and English dataset) and models, specifically in the context of healthy controls (HC) vs. PD classification, and regression tasks to predict demographic and articulation characteristics. Our study serves as a continuation of our prior work introduced in Klempir et al. [20], which analyzed wav2vec for classification, this paper incorporates additional cross-database, regression and feature importance experiments.

## 2 Related Work

Speech and voice characteristics of individuals with PD exhibit variations influenced by age and gender [21]. Early-onset PD, occurring before the age of 50, introduces an additional layer of complexity in the understanding of these nuances [22]. The precise prediction of both chronological and biological age in PD constitutes a clinically significant task. Recent machine learning studies have leveraged extensive datasets, such as the UK Biobank brain imaging data, to construct brain-age models and investigate various aging-related hypotheses [23, 24]. One approach involves calculating the brain-age delta by subtracting chronological age from the estimated brain age [25]. In the context of PD, Eickhoff et al. conducted a comprehensive analysis on two PD cohorts (de-novo and chronic), revealing a substantial increase in biological age - of approximately three years - compared to chronological age, a phenomenon evident even in de-novo patients. This age discrepancy significantly correlates with disease duration, as well as heightened cognitive and motor impairment [26]. The prediction of age based on speech features typically employs neural networks, demonstrating an efficient level of accuracy in classifying individuals into age groups. For instance, studies have reported testing accuracies as high as 85.68% [27], and classification errors by age group of less than 20% [28].

DL methods have exhibited their efficacy in extracting valuable features from voice and speech, particularly in the classification between those with neurological disorders and HC [29]. Convolutional Neural Networks (CNNs) stand out as a class of DL methods that surpass classical machine learning approaches in terms of performance. CNNs have achieved remarkable accuracy rates, often exceeding 99% [30, 31]. Various CNN architectures are commonly employed in these applications, with data typically transformed into the time-frequency domain to retain both temporal and frequency information. This transformation can involve spectrogram-based Short-Time Fourier Transform (STFT), as observed in the analysis of PD vowel signals [31]. Additionally, CNNs can operate on mel-scale spectrograms of hyperkinetic dysphonia recordings [30], or utilize continuous wavelet transform to model articulation impairments in patients with PD [32]. Furthermore, research by Vásquez-Correa et al. demonstrated that fine-tuning CNNs through transfer learning contributes to enhanced accuracy in classifying patients with PD across different languages [33].

Recently, representation learning (RL) technology has facilitated the transformation of raw biomedical data into compact and low-dimensional vectors, commonly referred to as embeddings [34]. These embedding methods have gained widespread adoption across various biomedical domains, including natural language processing (NLP), where they are utilized to represent clinical concepts derived from unstructured clinical notes, such as symptoms and lab test results [35]. Compression algorithms, similar to DL autoencoders, have demonstrated efficacy in constructing embedding vectors for classification tasks, particularly on out-of-distribution domains [36]. Furthermore, NLP embedding techniques have been used to generate concise and scalable features for virome data, offering insights into the importance of each sequence position in the resulting supervised model outputs [37]. In RL for speech, embeddings serve as fixed-size acoustic representations for speech sequences of varying lengths [38]. The application of speech self-supervised learning (SSL) has enabled the utilization of large datasets containing unlabeled speech signals, achieving remarkable performance on speech-related tasks with minimal annotated data.

These SSL embedding systems have been comprehensively benchmarked, considering factors such as model parameters and accuracy across various tasks [39]. For accessibility, pre-trained Automatic Speech Recognition (ASR) embedding models are readily available through the Hugging Face repository [40].

Pre-trained embedding relevant to the automatic assessment of PD include x-vectors [41], TRILLsson [42], HuBERT [43], and wav2vec [8, 9]. In a comparative study by Favaro et al., the aforementioned architectures were evaluated for PD detection in multi-lingual scenarios, marking the initial application of TRILLsson and HuBERT in experiments related to PD speech recognition [44]. There are existing studies that showed the capability of x-vectors for analyzing PD speech [45, 46]. The method of wav2vec embedding, which is available in two versions (wav2vec 1.0 [8] and the transformer-based wav2vec 2.0 [9]), represents a class of DL designed for self-supervised, high-performance speech processing. Fine-tuned wav2vec has proven efficient across various speech recognition tasks and languages [14]. The recent application of the transformer-based wav2vec 2.0 showcased its utility in developing speech-based age and gender prediction models, including cross-corpus evaluation, with significant improvements in recall compared to a classic modeling approach based on hand-crafted features [47]. Additionally, wav2vec 2.0 representations of speech were found to be more effective in distinguishing between PD and HC subjects compared to language representations, including word-embedding models [48]. As a pre-trained model, wav2vec shares the advantage with TRILLsson and x-vectors of being directly applicable without the need for further training, addressing the data-hungry nature common to many neural networks in the field.

In 2023, the exploration of cross-database classification between HC and individuals with PD has attracted considerable attention [20, 49]. Hires et al. conducted a study on the inter-dataset generalizability of both deep learning and shallow learning approaches, specifically focusing on sustained vowel phonation recordings. Their findings showed an excellent performance during model validation on the same dataset, with a drop in accuracy during external validation [49]. Our previous study focused on the application of wav2vec 1.0 on reading speech, which presented a leave-one-group-out classification between Italian and English languages, with a relatively minor drop in performance observed [20]. The relevance of cross-database classification was emphasized by Javanmardi et al. [12], which investigated individual wav2vec layers for PD detection. Their conclusions highlighted the need for further research to explore the generalizability of wav2vec features, particularly in cross-database scenarios. Furthermore, the study [49] found out that distinct sets of features contribute differently across various datasets and the same set of features is not universally shared across models for individual datasets.

The International Journal of Medical Informatics has introduced a comprehensive checklist for the (self)-assessment of medical AI studies [50], emphasizing the utmost significance of feature importance and interpretability when introducing new methods for real-world applications. In the domain of speech recognition, mel-frequency cepstral coefficients (MFCCs) are emerging as key features for assessing speech impairments in neurological diseases; however, their interpretability remains limited [51]. First experiments have been conducted to explain MFCCs and move closer to achieving interpretable MFCCs speech biomarkers [52]. The study by Favaro et al. highlights another relevant finding: models based on non-interpretable features (DL embedding methods) outperformed interpretable ones [44]. Interpretable feature-based models offer valuable insights into speech and language deterioration, while non-interpretable feature-based models can achieve higher detection accuracy.

The remote assessment of (patho)physiological parameters, encompassing variables such as body temperature, blood oxygenation, heart rate, movement, and speech, holds significant relevance in scenarios where direct contact is impractical or must be avoided for various reasons. The collection of speech data, accessible from virtually anyone with an audio-enabled device, presents an opportunity to remotely screen for PD, thereby promoting inclusivity and accessibility in neurological care [21]. In contemporary biomedical artificial intelligence (AI) systems, particularly multimodal ones, an abundance of distinctive features or signals is embedded within the dataset [53]. Speech and voice play an important role in numerous instances [54]. To get the full potential of these complex datasets, advanced AI techniques can be employed to align multimodal features onto a shared latent space. This approach enhances the precision of phenotype prediction for highly heterogeneous data spanning various modalities [55]. We anticipate that the mentioned AI systems can be improved by incorporating speech representations like wav2vec. However, it is essential to conduct an investigation into the wav2vec component, examining its capabilities to generalize across different datasets and tasks.

## 3 Methods

### 3.1 Datasets

#### 3.1.1 Participants Rhythmically Repeat Syllables /pa/

This classification dataset involved 30 male PD patients and 30 male age-matched HC Czech participants rhythmically repeating the syllable /pa/ [7]. The data, consisting of audio signals with a sampling frequency of 48 kHz, focused primarily on evaluating “pa” recordings, a standardized speech examination in PD.

#### 3.1.2 Italian Study by Dimauro et al

This dataset, used for both classification and regression tasks, stems from a study assessing speech intelligibility in PD conducted at the Università degli Studi di Bari, Italy [56]. The dataset used for HC vs. PD classification included 50 subjects (elderly HC: n = 22, PD: n = 28), with measurements of text readings (44.1 kHz) available for each individual. For the regression experiments, we also included a young HC group (young HC: n = 15), along with metadata for regression tasks such as age and the estimated number of characters read per second. In the HC group, individuals aged 60-77 years were included, consisting of 10 men and 12 women. None of these individuals reported any particular speech or language disorders. Regarding the PD group, patients aged 40-80 years were included, consisting of 19 men and 9 women. In the young group, individuals aged 19-29 years were included, consisting of 13 men and 2 women.

#### 3.1.3 English Dataset

Chosen for classification experiments, the English dataset “Mobile Device Voice Recordings at King’s College London (MDVR-KCL)” [57] comprised of 21 HC and 16 PD subjects who read aloud “The North Wind and the Sun.” Measurements of text readings were available for each participant, with a sampling frequency of 44.1 kHz. There is no available information regarding the distribution of gender and age.

Our evaluation contained three PD datasets for classification, previously studied in [7, 20], and two additional PD quantitative features (*age*, *characters per second*) for regression tasks based on data from the Italian study [56].

### 3.2 Signal Processing and Feature Extraction

#### 3.2.1 Naive Loud Regions Segmentation

Based on the Italian study data, we derived an additional parameter for the purposes of regression tasks, which we refer to as *loud region duration*. The preprocessing phase involved segmenting the signals into binary loud regions. As a basic baseline approach, the raw audio signal was segmented using a straightforward formula, creating a binary vector representing quiet and loud regions. Each data point in the series was encoded as 1 if its absolute value exceeded the mean absolute value of the signal, and 0 otherwise. Subsequently, the binary vector underwent averaging through a 10,000-sample window rolling average and was summed to generate a single numerical value for each recording.

#### 3.2.2 MFCCs Features Calculation

To establish baseline comparisons, we calculated the audio features for the recordings stored in WAV files using the Librosa Python library [58]. The recordings were resampled at 16 kHz. Subsequently, the waveform underwent processing with Librosa to compute 50 MFCCs, with each coefficient averaged over time (MFCC-mean). This computation resulted in a vector comprising 50 values.

#### 3.2.3 Wav2Vec Embedding and Features Calculation

To assess the generalization capability across databases and minimize the need for feature engineering, we employed wav2vec 1.0 [8], a speech model that accepts a float array corresponding to the raw waveform of the speech signal. All recordings were resampled at 16 kHz, a prerequisite for the wav2vec embedding used for feature extraction. In contrast to traditional signal processing methods, wav2vec is capable of learning a suitable representation of an audio signal directly from data, avoiding the necessity of manual feature extraction [20].

For our experiments, we utilized pretrained components of the wav2vec model, which maps raw audio to a latent feature representation. This model is pretrained in an unsupervised manner and has demonstrated advanced performance in speech recognition tasks with minimal fine-tuning. Specifically, we obtained a publicly available wav2vec-large model trained with the LibriSpeech training corpus, which contained 960 hours of 16 kHz English speech [59].

Due to the dynamic representation of the wav2vec embedding over time [20], we computed a derived 1-D static feature vector of 512 dimensions for each recording based on three different statistics (mean: wav2vec-mean, standard deviation: wav2vec-std, sum: wav2vec-sum) along the time axis. The output of this method served as the input to machine learning models for classification and regression. Similar approaches exist in literature to aggregate specific DL audio embeddings, as seen in TRILLsson and wav2vec 2.0 [44]. Since feature extraction from long recordings requires relatively complex computation in [44], the authors divided long recordings into 10-second segments. However, with wav2vec for this research, we found this segmentation to be unnecessary, allowing us to process the entire signal without the need for splitting it into sections.

In an extension of our previous research [20], this study enriches the previous results by incorporating wav2vec-std. Additionally, a derived feature vector of length 20 is calculated, incorporating 10 Principal Component Analysis (PCA) components of wav2vec context (wav2vec-mean) and 10 PCA components of spectral MFCCs characteristics (MFCC-mean) combined into a single vector. Ten components were selected to capture a minimum of 90% of the original variance (cumulative sum of explained variance) across datasets for both wav2vec-mean and MFCC-mean. We aim for a compact resulting vector length with an equal number of components for both input vectors.

### 3.3 Modeling and Statistical Methods

A high-level overview of the proposed machine learning methodology is illustrated in Fig.1.

**Fig. 1.**
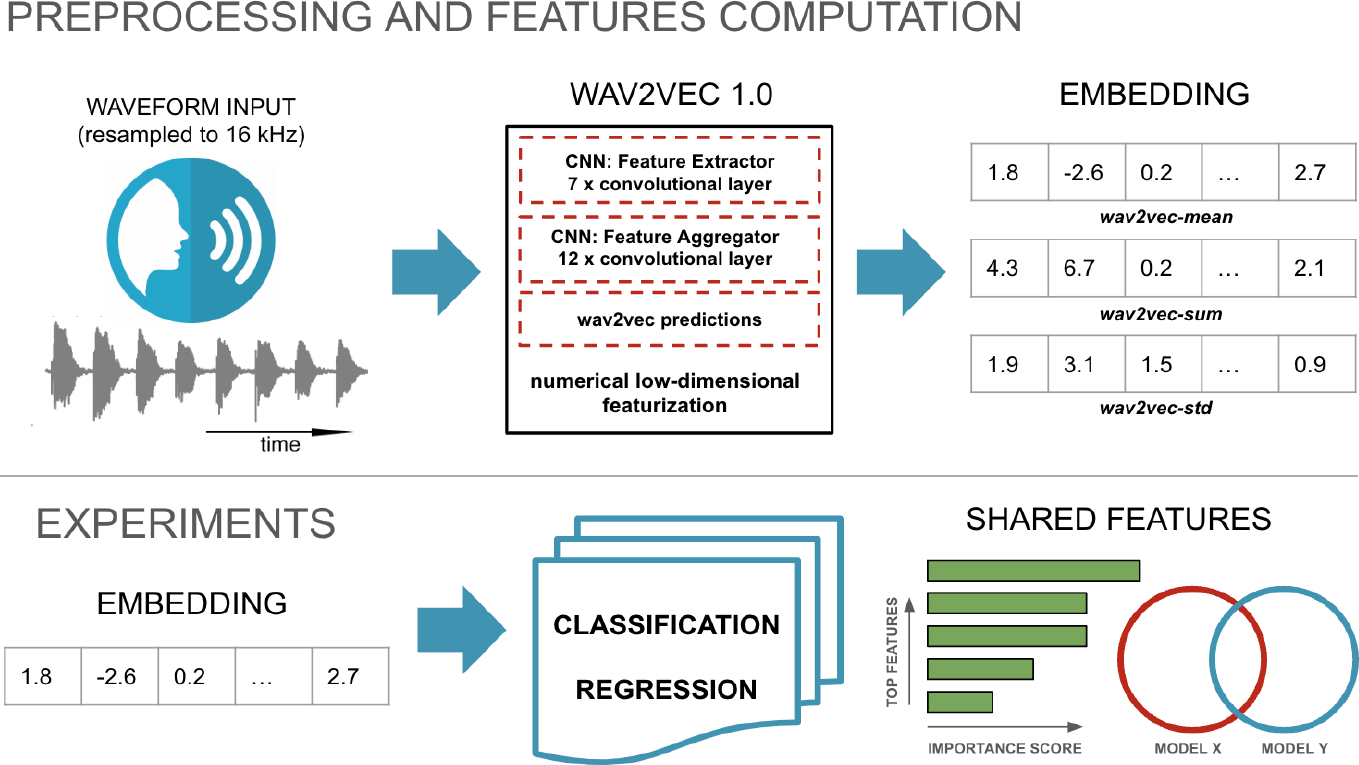
Illustrative diagram of the proposed signal processing and experiments.

#### 3.3.1 Cross-Database Classification Experiments

To assess the generalizability of wav2vec and MFCCs features across multiple PD detection datasets, we conducted a series of classification experiments. This experimental setup is entirely unique in this version of the paper compared to our prior publication [20]. Following a similar approach as recently presented in [49], we adopted combo scenarios. Specifically, we assessed performance on individual datasets, and more importantly, we implemented additional scenarios: one that utilizes a combination of multiple datasets for training and evaluates a completely unseen remaining dataset, and vice versa. For binary classification modeling, we employed an ensemble random forest classifier (with default settings, n_estimators=100 and criterion=’gini’) from the Python scikit-learn library. Model evaluation was conducted using 5-fold cross-validation with 5 repeated fits to measure the performance of the classification models. The interpretation of the classification results involved comparing the model output with the ground truth using a receiver operating characteristic (ROC), where the area under the curve (AUC) was employed to quantify the level of accuracy. In this study, the primary evaluation metric for model performance was the area under the ROC curve (AUROC).

#### 3.3.2 Regression Experiments

We utilized wav2vec features in regression modeling tasks for modeling age and parameters associated with articulation rate. In the modeling phase, we employed the lasso with alpha = 0.01 (least absolute shrinkage and selection operator) model from the Python scikit-learn library and conducted model evaluations using 5-fold cross-validation. Lasso employs regularization through a geometric sequence of Lambda values. Its primary advantage lies in the suppression of redundant features, contributing to improved model generalization. This characteristic proved particularly beneficial when dealing with a limited number of observations and a substantial number of features. To measure the performance of our model, the following metrics were computed: Spearman correlation coefficient (rho), r-squared (r2) metric and mean absolute error (MAE).

#### 3.3.3 Feature Importance and Common Features Across Tasks

To explore the hypothesis of a potential link between classification and regression tasks, we investigated the feature importance of the wav2vec embedding as an important aspect of interpretability. While wav2vec exhibits promising potential for applications in speech processing for neurological diseases, it is essential to grasp its behavior across various tasks, encompassing inter-task scenarios. The goal was to identify features that exhibit common features across tasks and evaluate their (dis)similarity. Approaches that involve models designed to handle irrelevant and unrelated features to the modeled variable, can lead to enhanced model explainability. Such examples are linear regression, where coefficients can be set to zero, or decision trees, which do not use irrelevant features in tree splitting criteria. The model choice not only improves model interpretability but also reduces computational requirements for training and model utilization. Our objective was to analyze global feature importance, which refers to the overall significance of a feature across all instances within the dataset. It’s essential to note that there is no universally superior method for determining global feature importance, as each method provides estimates based on distinct assumptions.

In the initial step, we identified the top 30 most contributing wav2vec features for each model considered. For classification, logistic regression, known for its feature importance computation built-in functionalities, was employed. For regression, we utilized the random forest regressor followed by the SHAP explainability method [60]. SHAP values were extracted from the best-fitting model for all individuals. In cases of more complex models, Shapley values are approximated through Monte Carlo sampling. The top features were obtained based on the importance of each factor, ranked from most to least important according to feature importance values. The calculation of feature importances for both built-in scikit-learn tools and SHAP typically involves utilizing the training set.

In the subsequent step, the resulting top 30 most important features for each model were then subjected to statistical testing to assess the significant coverage of common features shared between models. The Fisher exact test was used to compute p-values. The computation considered the numbers of (non)-overlapping features and the total number of features. P-values below 0.05 were considered statistically significant. Visualization using Venn diagrams were used to illustrate the unique/common sets of features observed among the models. As an alternative approach to evaluate significance, we conducted a simulation experiment to identify the minimum number of shared features considered significant. In 10,000 runs, two sets of 30 features were randomly generated from a pool of 512 total features, and their intersection was calculated. The mean number of shared features was 1.76, with 5 shared features (99th percentile) demonstrating statistical significance (associated with a p-value of 0.01). To address potential concerns related to multiple comparisons, we applied Bonferroni correction to establish the corrected threshold for significance. A significance threshold was maintained, with a minimum of 6 shared features remaining statistically significant after correction. This corresponded to a p-value of 0.013, obtained by multiplying the base p-value of 0.004 by a factor of 3 (reflecting the number of independent tests for each presented Venn diagram).

## 4 Results

### 4.1 Intra and Inter-Dataset Classification for Detection of PD

This section presents the outcomes of the four primary classification scenarios detailed in Table 1. The table reports the AUROC scores achieved by the respective models on individual datasets (intra-dataset classification) and on combined datasets with external evaluation (inter-dataset classification). A subset of the results discussed here was previously addressed in [20], and these results are appropriately marked in Table 1.

**Table 1.** The AUROC scores represent the classification performance of the respective models in distinguishing between HC and individuals with PD across four scenarios, including intra-dataset and inter-dataset comparisons; wav2vec is abbreviated to w2v. * results presented and discussed in [20] ** 10 PCA from w2v-mean + 10 PCA from MFCC-mean

| # | TRAIN | TEST | w2v-mean | w2v-sum | w2v-std | MFCC-mean | 10 PCA w2v-mean | 10 PCA MFCC-mean | MFCC-mean +w2v-mean** |
| --- | --- | --- | --- | --- | --- | --- | --- | --- | --- |
| 1 | pa | pa | 0.61* | 0.82* | 0.70 | 0.78 | 0.58 | 0.84 | 0.81 |
|  | Italian | Italian | 0.98* | 0.97* | 0.98 | 0.98 | 0.98 | 0.99 | 0.99 |
|  | Eng | Eng | 0.80 | 0.71 | 0.80 | 0.74 | 0.65 | 0.79 | 0.77 |
| 2 | Italian, Eng | Italian, Eng | 0.94 | 0.90 | 0.93 | 0.89 | 0.83 | 0.81 | 0.88 |
|  | Italian, pa | Italian, pa | 0.87 | 0.89 | 0.86 | 0.92 | 0.81 | 0.89 | 0.91 |
|  | Eng, pa | Eng, pa | 0.72 | 0.75 | 0.71 | 0.71 | 0.58 | 0.81 | 0.78 |
| 3 | pa | Italian | 0.46 | 0.50 | 0.50 | 0.68 | 0.68 | 0.40 | 0.57 |
|  | pa | Eng | 0.40 | 0.50 | 0.51 | 0.49 | 0.34 | 0.68 | 0.55 |
|  | Italian | pa | 0.48 | 0.43 | 0.46 | 0.49 | 0.61 | 0.63 | 0.68 |
|  | italian | Eng | 0.72* | 0.56 | 0.67 | 0.47 | 0.56 | 0.42 | 0.51 |
|  | Eng | Italian | 0.90* | 0.78 | 0.77 | 0.51 | 0.73 | 0.34 | 0.47 |
|  | Eng | pa | 0.46 | 0.53 | 0.48 | 0.44 | 0.48 | 0.60 | 0.60 |
| 4 | Italian, Eng | pa | 0.49 | 0.43 | 0.52 | 0.55 | 0.53 | 0.63 | 0.64 |
|  | Italian, pa | Eng | 0.72 | 0.61 | 0.61 | 0.56 | 0.46 | 0.46 | 0.51 |
|  | Eng, pa | Italian | 0.86 | 0.80 | 0.88 | 0.56 | 0.75 | 0.50 | 0.64 |

The first scenario involved models trained on individual datasets and evaluated on the same datasets, covering three cases—/pa/, Italian, and English datasets separately. For /pa/, the top-performing model was achieved with 10 PCA MFCC-mean, slightly outperforming the wav2vec-sum. The combination of wav2vec and MFCCs features did not enhance overall performance. Remarkably, an excellent performance was observed across all models for the Italian dataset. In the case of the English dataset, AUROCs varied, with 0.74 for the MFCC-mean model and the best-performing model reaching 0.80 for wav2vec-std.

The second scenario examined the performance of the three models on combined datasets, where two datasets were mixed to create a larger dataset. The aim was to increase the volume of training data, potentially enhancing model performance. When combining the Italian and English datasets, overall, wav2vec yielded superior results compared to MFCCs. It is noteworthy that the mixing of wav2vec and MFCCs PCA components elevated the AUROC to 0.88, surpassing the individual AUROC values of 0.83 for wav2vec-mean and 0.81 for MFCC-mean. Conversely, in the case of merging Italian and /pa/ datasets, MFCCs outperformed wav2vec parameters. Additionally, for the mixed case of English and /pa/ datasets, 10 PCA MFCC-mean exhibited a higher AUROC (by 5-10%) compared to other models.

Scenario 3 comprised the most extensive set of results. A total of six models were trained to assess cross-database generalizability. When trained on /pa/, wav2vec features failed in accurately classifying HC vs. PD in both the Italian and English datasets. Some non-random generalizability signals (AUROC = 0.68) were observed in the cases of MFCC-mean and 10 PCA wav2vec-mean in the Italian dataset. The same AUROC was achieved for the English dataset in the case of 10 PCA MFCC-mean. When trained on Italian data, the combination of PCA components of wav2vec and MFCCs features proved to be the most effective when evaluating /pa/. Evaluating the Italian model on the English dataset revealed that wav2vec, previously shown to be promising in detecting HC vs. PD, outperformed the newly introduced MFCC-based features. A similar trend was observed in the reverse case, i.e., training on English and evaluating on Italian. Lastly, evaluating the English model on /pa/ resulted in decreased accuracy.

In Scenario 4, all wav2vec models trained on mixed English and /pa/ datasets achieved an AUROC higher than 0.8. The minimum was 0.8 for wav2vec-sum, and the maximum was 0.88 for wav2vec-std. Otherwise, we noticed analogous trends as those observed in Scenario 3.

Overall, the selected cross-database results obtained on individual datasets or mixed datasets were highly satisfactory, indicating a non-random level of generalizability in language-independent models for discriminating between PD and HC.

### 4.2 Regression Models to Predict Age and Articulation

The next section provides an examination of the results obtained for patient characteristics from the Italian dataset. Initially, we explored paired correlations among *age*, *characters per second*, and *duration* parameters with naive loud region segmentation (Fig. 2a). Subsequently, we utilized wav2vec-mean and the features were reduced to two dimensions using PCA. The PCA visualizations (Fig. 2b) of the wav2vec-mean embedding illustrated the correlation structure with the parameters from Fig. 2a. Individual PCA plots, color-coded with the respective parameters values, revealed patterns where similar values clustered closer to each other, confirming that capturing the age and articulation parameters based on wav2vec is highly likely. Furthermore, we visually examined the impact of gender on age prediction by previewing 2 PCA components of the wav2vec-mean (Fig. 3).

**Fig. 2.**
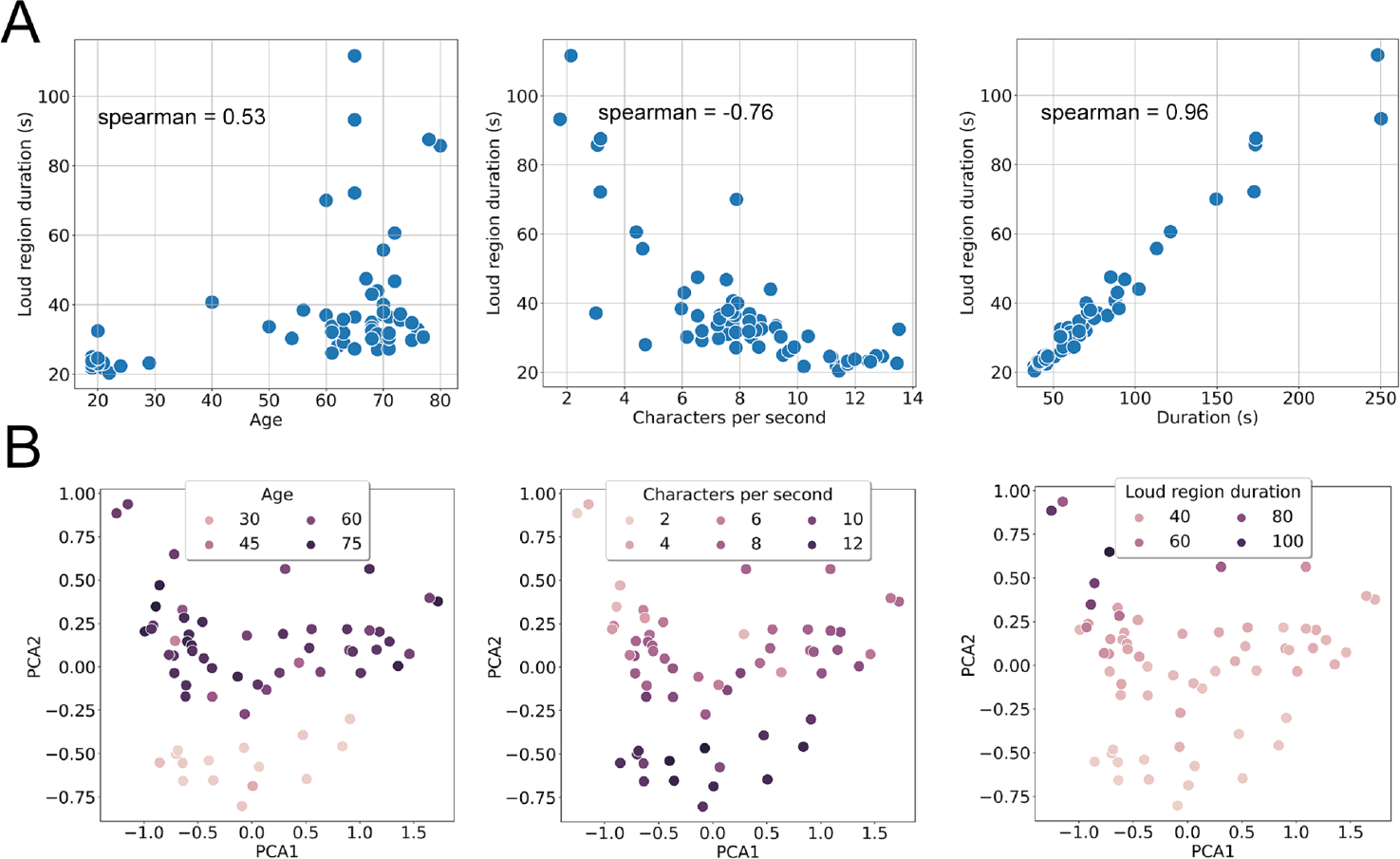
a) Correlations between the features from the Italian dataset and the calculated duration of loud regions. b) PCA plots depicting the relationship between wav2vec-mean features. The close grouping of similar colors suggests a nonrandom signal across all wav2vec-mean features. Each data point corresponds to one subject.

**Fig. 3.**
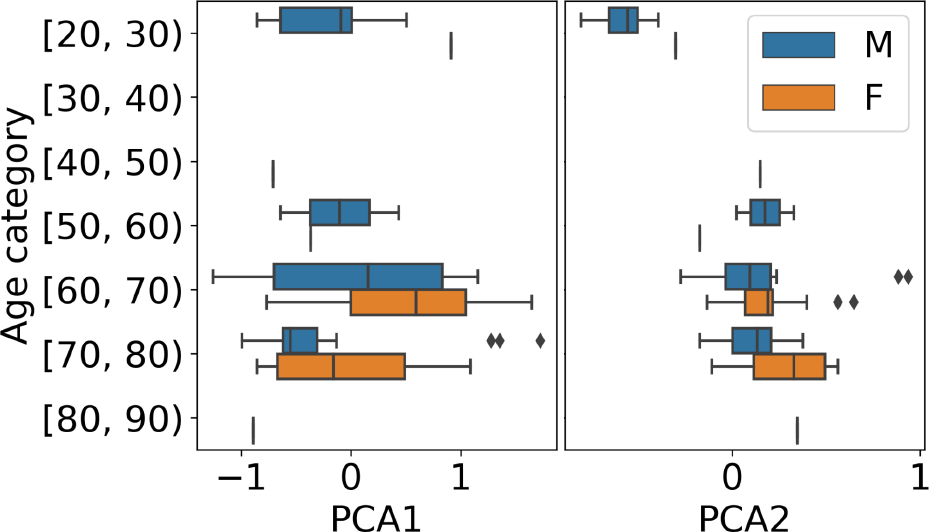
Boxplots illustrating 2 PCA components of wav2vec-mean, organized into distinct classes based on age and presented in separate panels for each component. The plots emphasize gender-related differences.

The trained wav2vec-mean regression models demonstrated correlations with *age* (Spearman R = 0.56), *characters per second* (Spearman R = 0.74), and *loud region duration* (Spearman R = 0.84). Additional metrics and the reported performance of the trained lasso regression models are presented in Fig. 4.

**Fig. 4.**
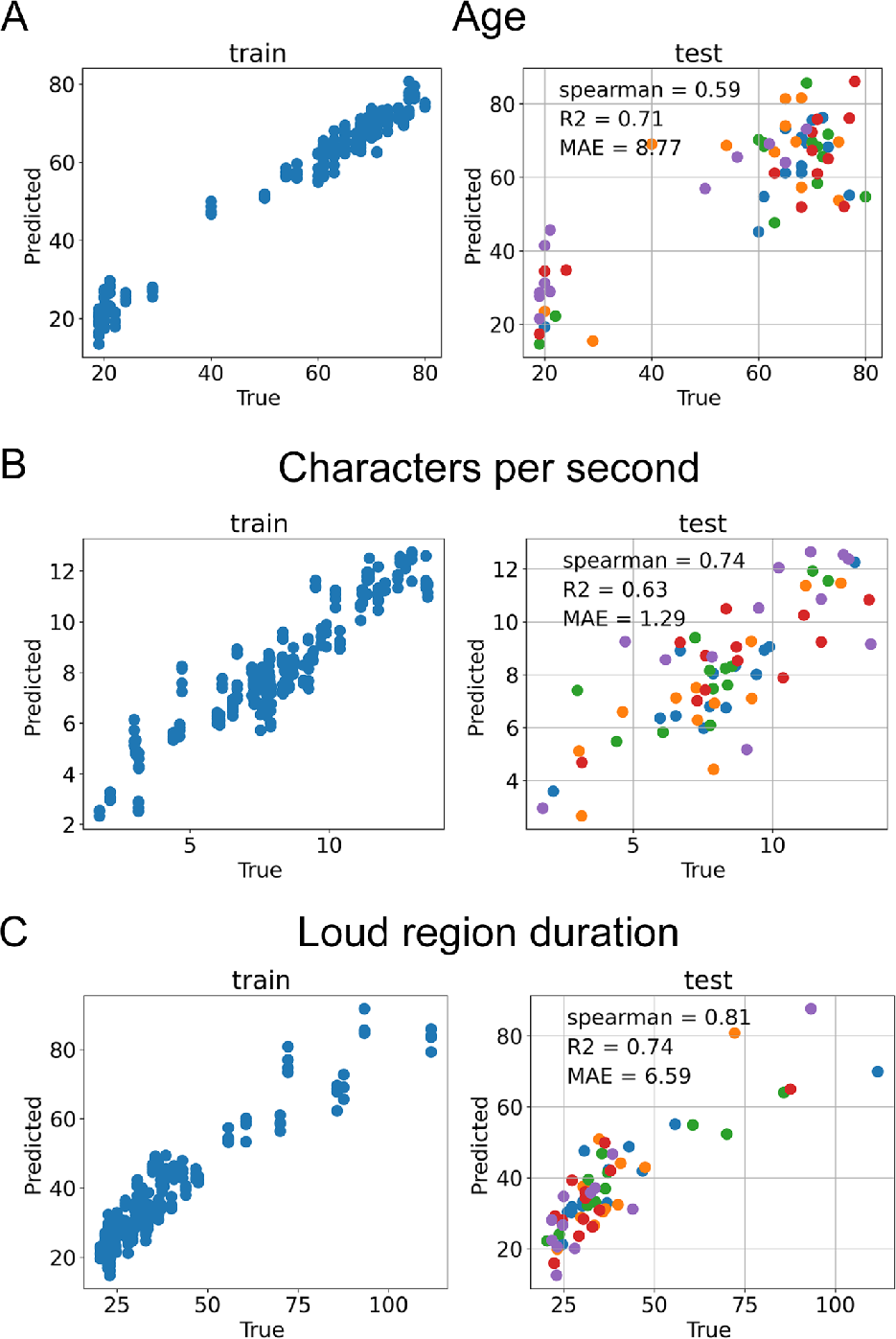
Performances of audio-wav2vec-based regression models on both training and testing data. Each distinct color in the plot corresponds to an individual run of cross-validation, specifically: a) modeling *Age*, b) modeling *Characters per second*, and c) modeling *Loud region duration*.

### 4.3 Exploration of Common Important Features Across Models

Feature importance assessments were conducted for various classification and regression models utilizing wav2vec-mean features. We focused on the top 30 impactful features, ranked from the most influential to the least, out of the complete set of 512 features. The connections between classification and regression model profiles are presented in Fig. 5a and Fig. 5b, respectively. Overall, the Fisher exact tests indicated a significant number of common features in all model pairs (p < 0.05). In the binary classification models (Fig. 5a), the strongest association was identified between Italian and English speech datasets (11 shared features). For regression (Fig. 5b), the overlap between the model explaining *loud region duration* and the model explaining *characters per second* was more pronounced (10 shared features) than with the model explaining *age* (5 shared features). The increased number of shared features between *loud region duration* and *characters per second* aligns with the observed correlation coefficients between *loud region duration* vs. *characters per second* (Spearman = -0.76), compared to the correlation between *loud region duration* vs. *age* (Spearman = 0.53), as shown in Fig. 2a. Taking into account the stringent Bonferroni corrected comparisons, certain pairs that had shown significance using the Fisher exact test were no longer significant (i.e. had fewer than 6 shared features). This applied specifically to the classification comparison between /pa/ and English, as well as both articulation rate models versus age.

**Fig. 5.**
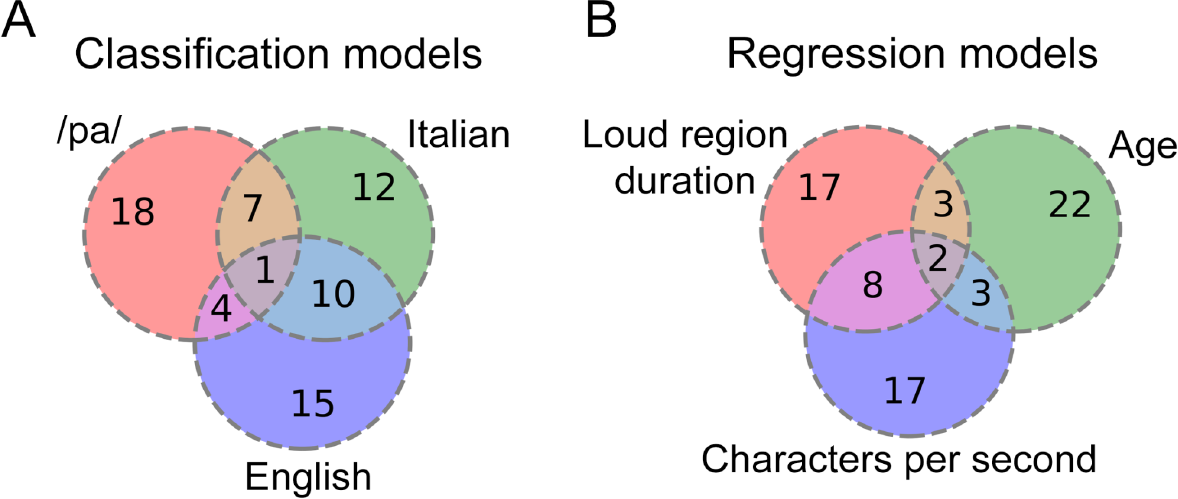
Venn diagrams illustrating shared features among multiple models: a) classification and b) regression.

Furthermore, we investigated the potential overlap between classification and regression tasks in the context of a binary *Age* classification model versus three regression tasks (Fig. 6). The parameter *Age* was considered in two distinct forms: firstly, as a binary class (young vs. elderly), and secondly, as a quantitative regression model predicting age. Concurrently, two additional models, addressing entirely different aspects, were examined and tested against the important features derived from the *Age* classification model. The association between the number of features in *Age:classificatio*n vs. *Age:regression* model was found to be significant, indicating shared features between the tasks. In contrast, other cases were not found to be statistically significant and over-represented.

**Fig. 6.**
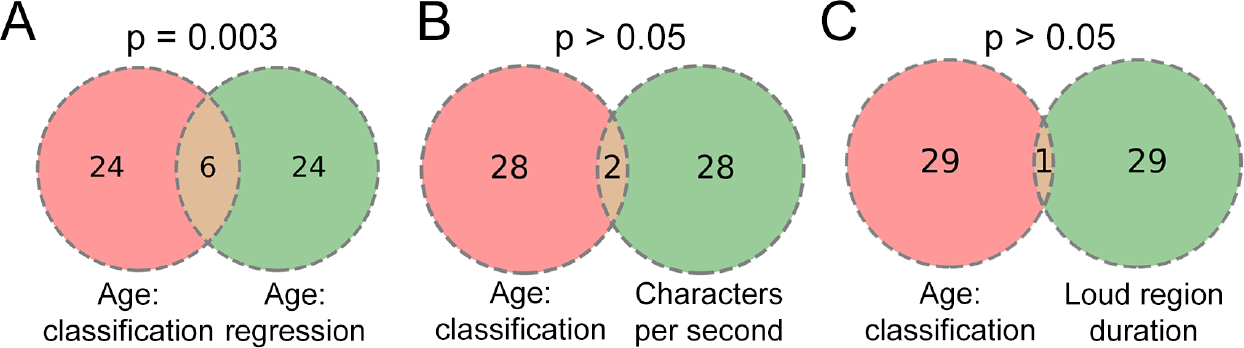
Comparative analysis of important features identified by the binary *Age* classification model vs. top features derived from three regression models.

## 5 Discussion

This paper introduces a study on using wav2vec 1.0 embeddings and MFCCs to detect Parkinson’s disease from speech, focusing on cross-corpora classification and regression tasks. It demonstrates wav2vec’s performance in PD detection over traditional methods and explores its generalizability across languages and tasks.

In addressing the primary hypothesis concerning the exploration of shared common features, our study focuses on the feasibility of achieving satisfactory results through cross-database classification. The main component is covered by machine learning, where we aimed to discern patterns and features that exhibited consistency across diverse databases. We constructed multiple models through various train-test combinations to evaluate the efficacy of wav2vec as a robust speech representation for PD detection. The main classification findings in Table 1 revealed that external validation did not result in a significant decrease in the AUROC metric for specific scenarios, importantly, when training on English and evaluating on Italian (AUROC = 0.90). This marked improvement stands in contrast to the baseline MFCCs features (AUROC = 0.51). Additionally, other new findings were observed. Due to the high correlation among the wav2vec embedding features, we performed a calculation of 10 PCA. This approach demonstrated superior results in some cases, outperforming models trained on the full list of features, especially in conjunction with baseline MFCCs and /pa/ dataset. We observed that wav2vec-mean excelled in the classification of read text, whereas a wav2vec-MFCC combination proved partially effective for tasks involving repeating syllables (/pa/). This behavior suggests that wav2vec is well-suited for external validation of complex speech tasks, while the combination of wav2vec and MFCCs performs better in simpler voice tasks. In conclusion, our classification experiments underscore the utility of wav2vec in cross-database PD detection.

In connection with existing peer-reviewed publications, our findings share relevance with the study conducted by Hires et al. [49]. The paper investigated the inter-dataset generalization of traditional and CNN machine learning approaches for PD detection based on voice recordings, specifically focusing on vowels. In their study, external validation demonstrated a decline in accuracy, with the highest AUC surpassing 0.7 in certain instances. Our research adopts a similar validation strategy, but we did not perform any hyperparameter tuning. In a recent study [44], the assessment of PD using wav2vec 2.0 and other DL embeddings in a cross-lingual context was explored. The investigation compared interpretable and non-interpretable features. Relevant to our results, the study’s cross-lingual experiments revealed significant performance variations based on the target language, emphasizing that DL embeddings can achieve an AUC exceeding 0.9 in cross-language validation. This finding supports our results obtained from wav2vec 1.0 and suggests possible alignment with other DL embeddings, such as TRILLsson. In examining the shared features between classification and regression tasks, our analysis of feature importances turned out that there isn’t a universally identical set of features shared across models for individual datasets. This variability is expected, especially when dealing with hundreds of features. Given the substantial number of features within wav2vec (the same applies to other DL embeddings as well), our focus was on the most impactful ones, extracting the top 30 features for each model. A strong cross-language signal pattern was observed between Italian and English speech classification models, indicating an enhanced level of generalizability across languages, specifically in the classification of PD versus healthy control—a trend consistent with our observed cross-database classification results. This finding aligns with existing research [61], demonstrating that unsupervised pretraining effectively transfers across languages. Identifying the most important features in DL embeddings holds the potential to interpret DL model outcomes by exploring cross-task relationships, enhance model performance, streamline model complexity by removing less critical features, mitigate overfitting, and further improve the generalization of PD detection from speech. In addition, in the context of our research, where we utilized MFCCs analysis as a baseline, Rahman et al. conducted a feature importance analysis on MFCCs [21]. Their investigation revealed that features that had an impact on the model’s performance were typically spectral features.

The findings of this study hold potential implications for broader applications in diverse contexts. As DL in PD continues to advance, overcoming technical and methodological challenges is crucial for widespread technology adoption. Sigcha et al. has highlighted the importance of large unsupervised data collection and the use of semi-supervised deep learning approaches to enhance PD symptom detection and severity estimation, ultimately improving the generalization capability of developed solutions [4]. The deployment of the wav2vec in a multimodal system, either separately as a one component, or in combination with MFCCs, offers opportunities for external validation and performance enhancement. For instance, incorporating Mel spectrograms for the audio component, as demonstrated in [54], can contribute to the development of multimodal biomedical AI. Additionally, the results obtained in remote speech PD, combined with a remote system like finger tapping [62], open possibilities for diverse applications at home. Moreover, utilizing federated learning for PD detection, locally extracted using the wav2vec 2.0 model, encourages further exploration of techniques for the generalization of models from databases of the same pathology, in different languages, without the need for sharing information between other institutions [63].

This paper has some limitations. First, our exploration solely focused on one wav2vec architecture, disregarding newer embeddings like wav2vec 2.0, which has demonstrated a slight improvement (2-3% in average recall) over wav2vec 1.0 in emotion recognition [11]. Although the difference between wav2vec versions is not substantial, there is potential for enhancement. Secondly, it is important to note that caution should be exercised in interpreting wav2vec representations in specific cases, such as slow or fast speakers, where model predictions may be less accurate. Further investigation and error analysis, involving additional datasets, are warranted for confirmation. Finally, the regression model for the age parameter in the Italian dataset reveals an apparent gap between young and elderly individuals, leaving some age ranges uncovered. This gap could introduce bias in the accuracy of the trained model, potentially leading to failures in predicting unseen data. Retraining the model separately for young and elderly groups might be necessary to address this limitation.

For future work, extending our experiments to include testing generalizability in other out-of-domain datasets, such as snoring [64], would be beneficial. Additionally, we anticipate further enhancements by incorporating proper augmentation techniques, such as audio-specific methods like noise addition or volume control, to introduce variability to the recordings and perturb the models. Initial attempts related to wav2vec augmentation have been explored in [65], and a recent study has provided a comparison of data augmentation methods in voice pathology detection [66]. Furthermore, the role of augmentation in wav2vec model training itself (wav2vec-aug) has demonstrated a substantial improvement in accuracy [67].

## 6 Conclusion

Based on the experimental results, this study showed the suitability of wav2vec for accurate intra-dataset classification and satisfactory performance in inter-dataset classification. It has proven effective in diverse regression tasks, such as modeling *age* and articulation characteristics like *characters per second* or *loud region duration*. Of utmost significance, the analysis of global feature importances reveals shared features among classification datasets, regression models, and statistically significant link between classification and regression tasks. While it cannot be definitively concluded that only a small subset of the 512 wav2vec features would universally solve any classification or regression task, the observed pattern suggests that similar tasks share common features. Transferability was more pronounced for similar speech tasks, such as reading or spoken text, evident in both shared feature importances and cross-database classification. The concluding insights emphasize the importance of feature importance methods in interpreting the generalization capability of developed deep learning solutions. The study proposes wav2vec embedding as a promising step toward a speech-based universal model to assist in PD evaluation.

## Data Availability

The Italian and English datasets analyzed during the current study are publicly available [56, 57]. Italian data [56] is available at the following URL: https://ieee-dataport.org/open-access/italian-parkinsons-voice-and-speech. English data [57] is available at the following URL: https://zenodo.org/records/2867216. The dataset comprised of participants rhythmically repeating the syllable /pa/ is not openly available and is available from the corresponding author upon reasonable request.

## Acknowledgements

We thank David Prihoda for the stimulating discussions about the wav2vec method and for his comments on earlier versions of the manuscript. This work was supported by the project of the National Institute for Neurological Research (Programme EXCELES, ID Project No. LX22NPO5107) - funded by the European Union – Next Generation EU.

